# Phylogenetic context of antibiotic resistance provides insights into the dynamics of resistance emergence and spread

**DOI:** 10.1101/2025.06.04.25328982

**Authors:** Kyle J. Gontjes, Aryan Singh, Sarah E. Sansom, James D. Boyko, Stephen A. Smith, Ebbing Lautenbach, Evan Snitkin

## Abstract

**Background:** To ameliorate the antibiotic resistance crisis, the drivers of resistance emergence (i.e., *de novo* evolution) and resistance spread (i.e., cross-transmission) must be better understood.

**Methods:** Whole-genome sequencing and susceptibility testing were performed on clinical carbapenem-resistant *Klebsiella pneumoniae* isolates collected from August 2014 to July 2015 across 12 hospitals. Ancestral state reconstruction partitioned patients with resistant strains into those that likely acquired resistance via *de novo* evolution or cross-transmission. Logistic regression was used to evaluate the associations between patient characteristics/exposures and these two pathways: resistance due to predicted within-host emergence of resistance, and resistance due to predicted cross-transmission. This framework is available in the user-friendly R package, *phyloAMR* (https://github.com/kylegontjes/phyloAMR).

**Results:** Phylogenetic analysis of 386 epidemic lineage carbapenem-resistant *Klebsiella pneumoniae* sequence type 258 isolates revealed differences in the relative contribution of *de novo* evolution and cross-transmission to the burden of resistance to five antibiotics. Clade-specific variations in rates of resistance emergence and their frequency and magnitude of spread were detected for each antibiotic. Phylogenetically-informed regression modeling identified distinct clinical risk factors associated with each pathway. Exposure to the cognate antibiotic was an independent risk factor for resistance emergence (trimethoprim-sulfamethoxazole, colistin, and beta-lactam/beta-lactamase inhibitors) and resistance spread (trimethoprim-sulfamethoxazole, amikacin, and colistin). In addition to antibiotic exposures, comorbidities (e.g., stage IV+ decubitus ulcers) and indwelling devices (e.g., gastrostomy tubes) were detected as unique risk factors for resistance spread.

**Conclusions:** Phylogenetic contextualization generated insights and hypotheses into how bacterial genetic background, patient characteristics, and clinical practices influence the emergence and spread of antibiotic resistance.

## INTRODUCTION

Antibiotic resistance is a significant public health challenge [1]. The development of infections with antibiotic-resistant pathogens significantly reduces treatment options and clinical cure rates. Of greatest concern are multidrug-resistant lineages, such as carbapenem-resistant *Klebsiella pneumoniae* (CRKP) sequence type 258 (ST258), that have evolved resistance to multiple classes of antibiotics and have disseminated worldwide [2].

The increasing problem of antibiotic resistance demands urgent clarity on the precise drivers of resistance proliferation, particularly in healthcare settings where the most concerning multidrug-resistant strains are commonly observed. To this end, previous studies have sought to identify patient risk factors associated with harboring a pathogen that exhibits antibiotic resistance [3–5]. However, this approach has a significant limitation - it overlooks the distinct routes to the acquisition of resistance, notably *de novo* evolution (i.e., the emergence of a unique resistant strain spontaneously in an individual) and cross-transmission of circulating resistant lineages (i.e., the acquisition of a resistant strain that is epidemiologically linked to other individuals) [6]. These two pathways call for distinct intervention strategies. For example, judicious antimicrobial stewardship may be useful to reduce selection for *de novo* resistance evolution, whereas measures to minimize cross-transmission include patient cohorting, hand hygiene compliance, and contact isolation [7–10]. A deeper understanding of these pathways to resistance acquisition will facilitate the application of tailored prevention strategies and sharpen our comprehension of how bacterial genetic background, patient characteristics, and medical practices drive the emergence and spread of antibiotic resistance.

Here, we developed an approach that uses phylogenetic context to partition antibiotic-resistant isolates into those putatively derived from *de novo* evolution versus cross-transmission. We then applied this method to understand drivers of these two pathways to resistance in the epidemic CRKP ST258 lineage, using a comprehensive collection of clinical isolates and patient metadata collected over one year from a network of long-term acute care hospitals (LTACHs). Both the emergence and spread of five critical antibiotic resistance phenotypes were identified, with the CRKP ST258 sub-lineage, antibiotic exposures, and patient clinical characteristics being differentially associated with the emergence and spread of each resistance phenotype.

## METHODS

### Sample collection

Clinical CRKP isolates were obtained from August 1, 2014, to July 25, 2015, at 12 southern California LTACHs [11]. Bacterial isolates from blood, respiratory, urine, and wound cultures identified as *K. pneumoniae* by the regional microbiology laboratory were tested for phenotypic carbapenem resistance using the 2015 Centers for Disease Control and Prevention criteria [11].

### Clinical metadata collection

Clinical metadata were extracted from the healthcare network’s electronic medical records, as described previously [11]. Patient demographics, medical comorbidities, and indwelling medical devices were recorded at specimen collection, along with antibiotic exposures up to 30 days prior to specimen collection. Antibiotic exposure data were available only during a patient’s stay in the LTACH.

### Antibiotic susceptibility testing to measure phenotypic resistance

Antibiotic susceptibility testing was performed for select aminoglycosides (i.e., amikacin, gentamicin), colistin, trimethoprim-sulfamethoxazole (TMP-SMX), and two recently approved β-lactam/β-lactamase inhibitor combinations (BL/BLI) (i.e., imipenem-relebactam, meropenem-vaborbactam) that were not clinically available at the time of specimen collection. Susceptibility profiles for TMP-SMX and gentamicin were extracted from clinical microbiology laboratory records. In contrast, susceptibility to amikacin, colistin, imipenem-relebactam, and meropenem-vaborbactam was determined using a custom broth microdilution panel (SENSITITRE, Thermo Scientific) [12]. Antibiotic resistance was defined as a minimum-inhibitory concentration within the intermediate or resistant category by the 2021 Clinical and Laboratory Standards Institute interpretative criteria for all antibiotics except TMP-SMX [12]., for which resistance was defined as a MIC ≥ 16/80 µg/mL (**Supplemental Table 1**). Resistance to BL/BLI agents was determined as a composite outcome of resistance to imipenem-relebactam and/or meropenem-vaborbactam.

### Whole-genome sequencing and phylogenetic tree reconstruction

Whole-genome sequencing of bacterial isolates was performed, as described previously [11]. Briefly, deoxyribonucleic acid was extracted using the DNeasy Blood & Tissue kit (QIAGEN). Short-read sequencing was performed on the Illumina HiSeq2500 instrument at the Pennsylvania Children’s Hospital of Philadelphia Microbiome Center. Sequencing quality was assessed using FastQC [13]. Trimmomatic trimmed adapter sequences and low-quality bases [14]. Variant calling was performed using SNPKIT v2.0 (https://github.com/Snitkin-Lab-Umich/snpkit) using the *K. pneumoniae* KPNIH1 reference genome (GenBank accession: <u>GCF_000281535.2</u>) [15].

The maximum-likelihood phylogenetic tree was reconstructed using a custom, in-house Snakemake pipeline [16, 17]. First, Gubbins identified recombination regions within the whole-genome alignment [18]. Phage regions identified by PHASTEST and repeat regions identified by nucmer were masked [19, 20]. The initial phylogenetic tree was reconstructed using FastTree’s Jukes-Cantor algorithm [21]. Subsequently, IQ-TREE was employed to determine the GTR+F+ASC model as the suitable model for use during Gubbins’ final round of tree reconstruction, which was performed using RAxML [22, 23]. Next, each isolate’s Gubbins-identified recombination regions were masked. Following this, IQ-TREE version 2.4.0 was executed using the GTR+F+ASC+R2 substitution model, which was inferred as the best model by their ModelFinder Plus algorithm [22, 24]. Default parameters were used, except that a total of 5,000 bootstraps and 500 unsuccessful iterations were required to halt the reconstruction. A single, closely related non-ST258 genome from this sample collection (BioProject accession, <u>PRJNA415194</u>) was used as the outgroup. Given the identification of numerous clades within our phylogeny, midpoint rooting was performed using the *phytools* R package to reflect the two bifurcating clades of ST258 accurately [25, 26].

### Phylogenetic analysis of antibiotic resistance using joint ancestral state reconstruction

We developed the open-source R package, *phyloAMR*, to study the emergence and spread of antibiotic resistance (https://github.com/kylegontjes/phyloAMR) [27].

PhyloAMR’s core function, *asr*, leverages *corHMM*’s ancestral state reconstruction algorithm to characterize the gain, loss, and continuation of antibiotic resistance across a phylogenetic tree [28]. Several alternative models were fit to the data and the rate matrix; the model with the lowest sample-size corrected Akaike information criterion (AICc) used for joint ancestral state reconstruction. Using inferred ancestral states and tip-based data, edges on the phylogenetic tree were evaluated to determine episodes where the trait continued (i.e., susceptible -> susceptible or resistant -> resistant), was gained (i.e., susceptible -> resistant), or was lost (i.e., resistant -> susceptible). This edge matrix can be leveraged for numerous applications, including characterizing the frequency of trait transitions across a phylogeny and investigating phenotype-genotype associations.

Here, we developed a phylogenetic tree traversal algorithm, *asr_cluster_detection*, that traces the ancestral states of each isolate to infer their evolutionary history. This algorithm takes an isolate’s trait data and walks upward on the tree to classify trait-containing isolates as phylogenetic singletons (i.e., evidence of *de novo* evolution of a trait) or members of a phylogenetic cluster of the trait (i.e., evidence that the trait was inherited from a common ancestor of circulating trait-containing lineage). Extending to the study of antibiotic resistance, resistant isolates with gain events inferred at the tip were classified as phylogenetic singletons.

However, instances where a resistant isolate had a gain event at the tip and a reversion event at its parental node were eligible for classification as members of a phylogenetic cluster. Resistant isolates were classified as members of a phylogenetic cluster if their ancestral gain event was shared with at least one additional resistant isolate. Resistant isolates that did not share an ancestral gain event with any other resistant isolate were classified as phylogenetic singletons. Phylogenetic clusters where all isolates belonged to one patient were not considered clusters and were reclassified as redundant phylogenetic singletons.

To describe the evolutionary history of antibiotic resistance, we characterized both the transitional data and the phylogenetic clustering of each antibiotic phenotype. The frequency of trait gain, loss, and continuation events was characterized using *phyoAMR*’s *asr_transition_analysis* function. Descriptive statistics for phylogenetic clustering were generated using *phyloAMR*’s *asr_cluster_analysis* function. Specifically, the crude frequency of the trait, descriptive statistics on the number of singletons, clusters, and summary statistics for cluster size. Two measures were employed to characterize the phylogenetics of a trait: phylogenetic occurrence (**Equation 1**) and clustering (**Equation 2**).

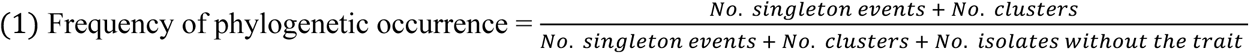

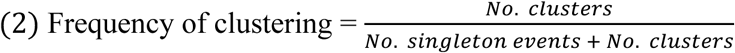

### Testing for heterogeneous rates of antibiotic resistance across clades

To further evaluate the influence of genetic background on the evolution of antibiotic resistance, we tested whether differences in rates of resistance evolution exist across the two clades of ST258. To achieve this, we utilized *phytool*’s *fitmultiMK* function, which implements a modified Markov model that allows for heterogeneous rates of discrete character evolution across user-specified regions on a phylogenetic tree [25, 29]. For each phenotype, we constructed a *fitmultiMK* model with a single regime, indicative of uniform rates of resistance evolution across the phylogeny. Next, a *fitmultiMK* model was constructed with two regimes painted on the tree, permitting each clade to have distinct rates of resistance evolution. Both the equal rates (ER) and all rates different (ARD) transition matrices were used for this analysis. The fit of the single-and multi-regime models was compared using the likelihood ratio test.

Next, hidden-rate modeling using *corHMM*, a data-driven approach, was performed to validate the presence of two distinct rate classes across the phylogeny. For each phenotype, ER and ARD models with one-and two-rate categories were constructed. Support for the existence of two rate classes was determined by comparing the AICc of the models.

### Regression modeling to identify risk factors associated with resistance emergence and spread

Logistic regression modeling was performed to evaluate the association between genetic background, patient demographics, and clinical variables with the primary resistance outcomes of crude resistance and our two phylogenetically-informed classifications of resistance. For each phenotype, patients with resistant isolates were partitioned as acquiring resistance via (1) putative *in-vivo* emergence (i.e., phylogenetic singletons) or (2) the acquisition of a strain belonging to a resistant lineage (i.e., members of a phylogenetic cluster), presumably due to a cross-transmission event, using the aforementioned methods. For patients with more than one isolate, their first isolate was selected unless they contributed a later resistant isolate, in which case the resistant isolate was chosen. Patients contributing only susceptible isolates served as the reference group for each analysis.

Logistic regression was first performed to evaluate the association between our resistance outcomes and three explanatory variables: an exposure history to the resistance phenotype’s cognate antibiotic, exposure to dysbiotic antibiotics, and having an isolate belonging to clade I of ST258. Exposure to dysbiotic agents-drugs that increase the risk for disruption of the gut microbiota and *Clostridioides difficile* infection-was defined as having a history of exposure to at least one of the following antibiotics in the preceding 30 days: cefepime, ceftaroline, ceftazidime, ceftriaxone, ciprofloxacin, ertapenem, metronidazole, levofloxacin, meropenem, imipenem, and piperacillin/tazobactam [30, 31]. Antibiotic exposure history was binarized as the receipt of ≥1 day of therapy in the past 30 days for that antibiotic. Regression models evaluating the association between our resistance outcomes and antimicrobial exposures were adjusted for age and sex.

Next, we performed a data-driven, multivariable regression modeling to identify shared and unique risk factors for our resistance outcomes. Eligible variables included patient demographics, clinical comorbidities, indwelling medical devices, and antibiotic exposure histories (**Supplemental Table 2**). The purposeful selection algorithm was implemented with minor alterations [32]. First, unadjusted logistic regression was performed. Next, all variables with an unadjusted p-value < 0.20 were included in a logistic regression model. An iterative process removed variables if their p-value was > 0.10. After compiling this model, all initially ineligible variables were iteratively included in the model and retained if they were identified their p-value was < 0.10.

### Data analysis and visualization

All data analysis and visualization, unless otherwise stated, were performed using R version 4.5.0 and corresponding *tidyverse* packages [33, 34]. The *ggplot2* and *ggnewscale* packages were used to generate common figures and scales [34, 35]. The *ape*, *phytools*, and *ggtree* packages were used to generate phylogenetic visualizations [25, 36, 37]. Descriptive tables were created using the *tableone* package [38]. Forest plots were generated using the *forestplot* package [39]. Finally, *cowplot* was used to generate multi-panel figures [40].

Code for this project can be found at https://github.com/kylegontjes/phylogenetic-resistance-ms/.

## RESULTS

### Study population

A total of 386 clinical CRKP ST258 isolates were collected from 312 patients across 12 California LTACHs over nearly 12 months (**Figure 1A-B**). Fifty-five patients (17.6%) contributed more than one CRKP ST258 isolate during the study period (**Figure 1C**). On first isolate collection, the median age was 72.7 years (interquartile range (IQR), 18.2 years), and 149 (47.8%) were female. Most patients had at least one indwelling medical device (87.8%), multiple medical comorbidities (64.4%), and received antibiotics in the 30 days preceding isolate collection (72.8%). The median LTACH length of stay before clinical culture was 21 days (IQR, 37 days). Cohort characteristics are provided in **Supplemental Table 3.**

**Figure 1.**
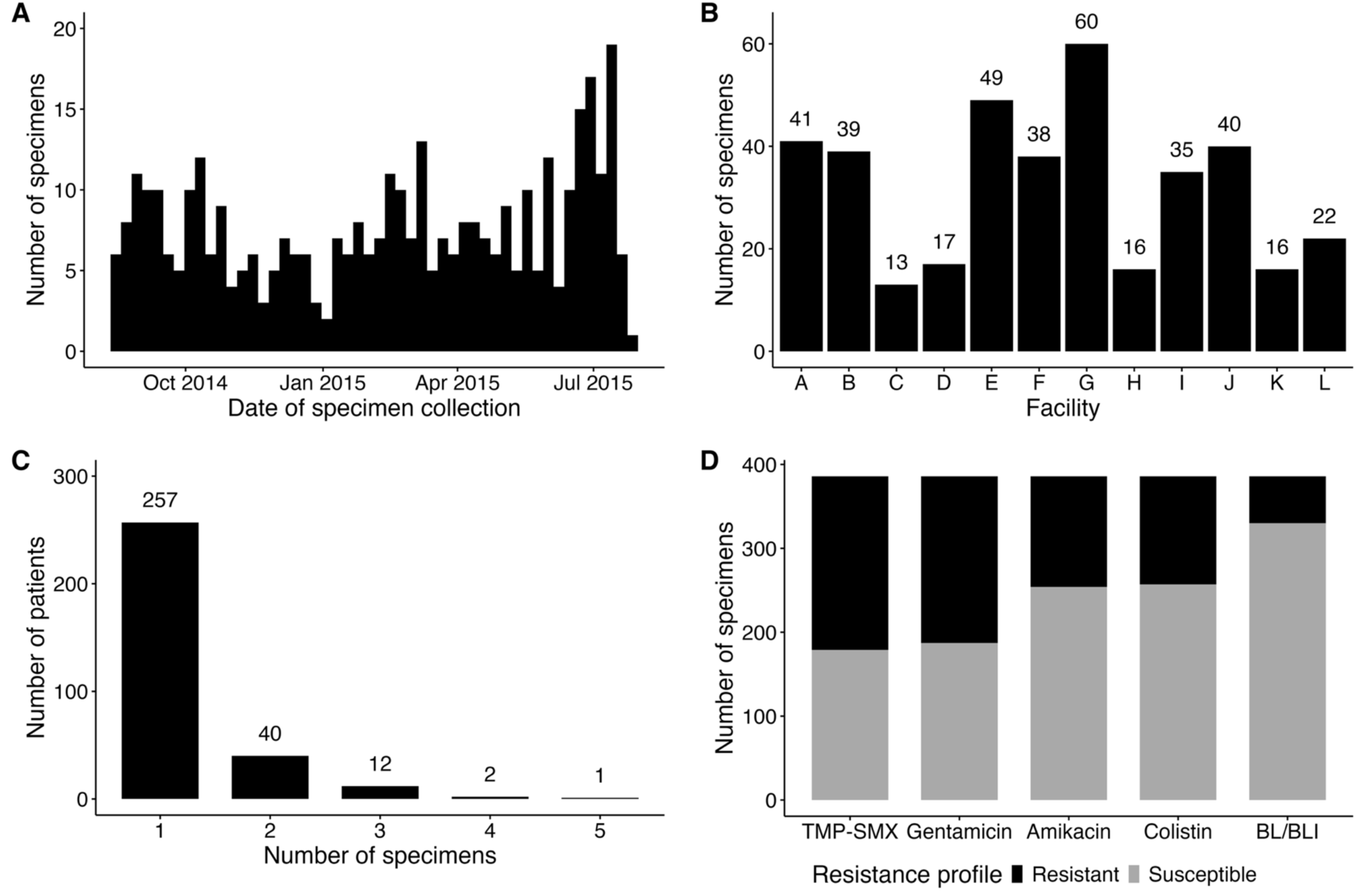
Comprehensive sampling of carbapenem-resistant *Klebsiella pneumoniae* that are resistant to last-resort or recently introduced antibiotics **Abbreviations:** BL/BLI, beta-lactam/beta-lactamase inhibitor; TMP-SMX, Trimethoprim-Sulfamethoxazole (A) Number of clinical carbapenem-resistant *Klebsiella pneumoniae* sequence type 258 isolates collected from August 1, 2014, to July 25, 2015. (B) Number of specimens collected across each of the twelve California long-term acute care hospitals. (C) Number of specimens contributed by each patient. (D) Number of resistant and susceptible isolates for five resistance phenotypes. Antibiotic resistance was defined as having a minimum-inhibitory concentration in either the resistant or intermediate category, per the 2021 Clinical and Laboratory Standards Institute criteria for all agents, except TMP-SMX, for which resistance was defined as MIC ≥ 16/80 µg/mL. Resistance to BL/BLI agents was determined as a composite of resistance to either imipenem-relebactam and/or meropenem-vaborbactam.

### Antibiotic susceptibility profiles of clinical carbapenem-resistant *Klebsiella pneumoniae* isolates

On application of clinical breakpoints, resistance was observed in 207 (53.6%) to TMP-SMX, 199 (51.6%) to gentamicin, 132 (34.2%) to amikacin, 129 (33.6%) to colistin, and 56 (14.5%) to novel BL/BLI combinations (meropenem-vaborbactam, 39 [10.1%]; imipenem-relebactam, 36 [9.3%]) (**Figure 1D)**. Minimum-inhibitory concentration distributions are reported in **Supplemental Figure 1**.

### Whole-genome sequencing revealed variable phylogenetic clustering of resistance

We implemented a phylogenetic tree traversal algorithm to characterize the emergence and spread of antibiotic resistance in this densely sampled population (**Figure 2A**). Overlaying resistance on the phylogenetic tree revealed numerous independent episodes of resistance emergence and spread for each antibiotic (**Figure 2B**). Quantification using our ancestral state reconstruction-informed algorithm revealed differences in the number and size of resistant clusters for different antibiotics (**Figure 2C-E**), suggesting differential propensities for resistance to emerge and spread. Indeed, ancestral state reconstruction produced evolutionary models with evolutionary rates that varied in magnitude (**Supplemental Table 4**), further supporting differences in emergence rates for different antibiotics. The ancestral states and phylogenetic clustering results for each phenotype were overlaid on the phylogenetic tree in **Supplemental Figure 2**.

**Figure 2.**
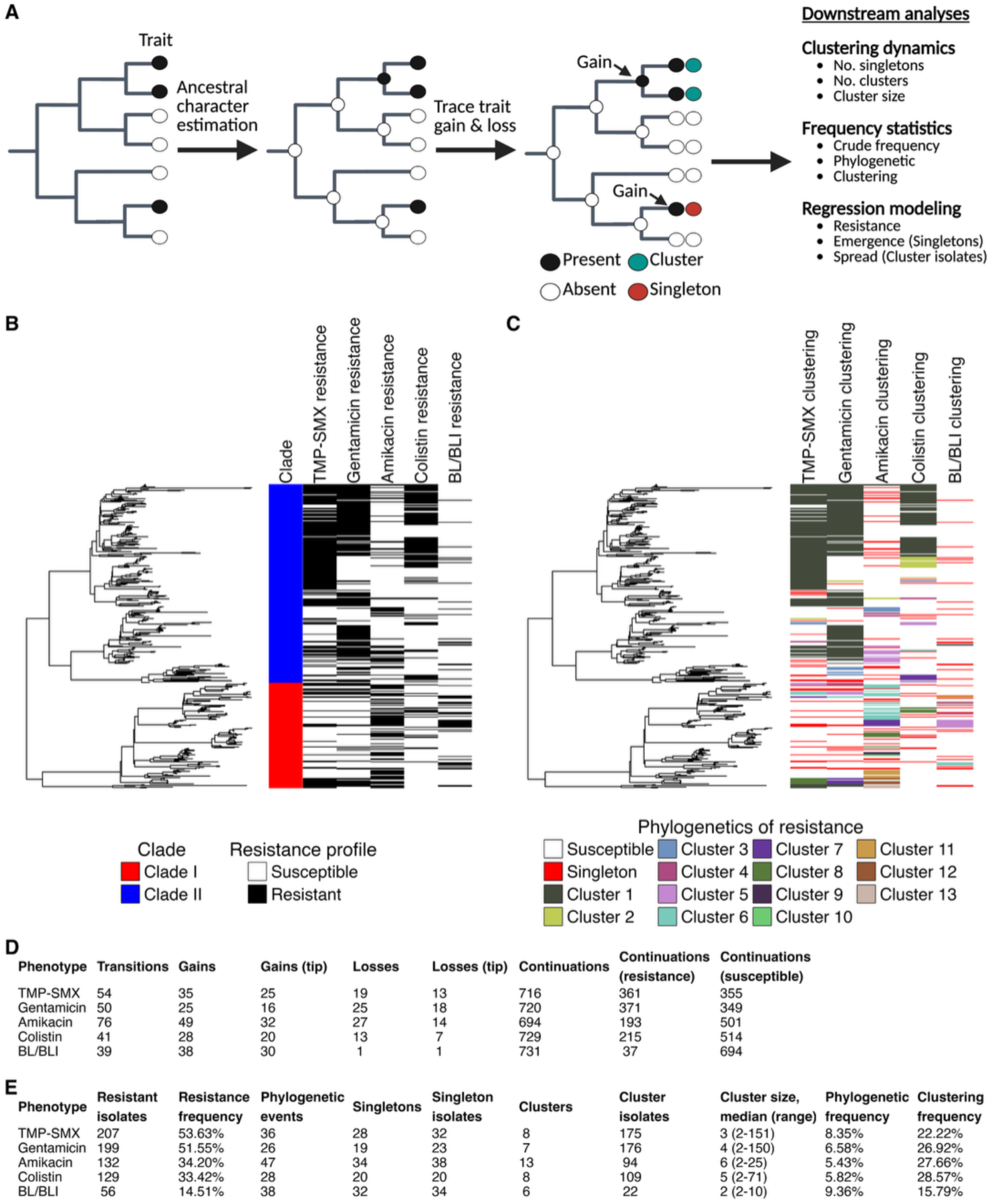
Phylogenetic characterization of resistance revealed differential patterns of resistance emergence and spread for five antibiotic phenotypes among clinical carbapenem-resistant *Klebsiella pneumoniae* sequence type 258 isolates **Abbreviations:** BL/BLI, beta-lactam/beta-lactamase inhibitor; NS, resistance; TMP-SMX, Trimethoprim-Sulfamethoxazole (A) Ancestral character estimation was performed to characterize the phylogenetic clustering of antibiotic resistance. Phenotypes were binarized into susceptible and resistant using clinical breakpoints. Resistance was inferred at internal nodes using *corHMM*’s joint ancestral reconstruction algorithm. The phylogenetic tree was traversed from parent to child node to determine episodes of trait continuation (i.e., susceptible -> susceptible and resistant -> resistant), gain events (e.g., susceptible -> resistant), and loss events (i.e., resistant -> susceptible). Resistant isolates were inferred as phylogenetic singletons if the resistant gain event was inferred at the tip with no history of susceptible reversion at their parental node or the resistant isolate did not share its gain event with another isolate. Resistant isolates were classified as members of a phylogenetic cluster if their gain event was shared with at least one additional resistant isolate. Lineages where all resistant isolates belonged to one patient were reclassified as redundant resistant singletons. Panel A was generated using BioRender. (B) Resistance overlaid across the *Klebsiella pneumoniae* sequence type 258 phylogeny. Black indicates resistance to the antibiotic, whereas white indicates susceptibility. (C) Phylogenetic clustering of resistance was overlaid across the phylogenetic tree. Red indicates resistant singletons, whereas the remaining colors indicate phylogenetic clusters of resistant isolates, as inferred from the ancestral state reconstruction algorithm. (D) Phylogenetic transition and (E) clustering statistics for each antibiotic. The frequency of phylogenetic occurrence accounts for the number of episodes a trait occurs across the phylogenetic tree relative to the total number of possible events. The frequency of clustering characterizes the proportion of phylogenetic events that are phylogenetic clusters.

In addition to differences across antibiotics, we also observed differences in phylogenetic clustering between the two major ST258 clades (**Figure 3A & Supplemental Table 5**). Testing for heterogeneity in evolutionary rates supported a role for the genetic background of ST258 in shaping the emergence and spread of antibacterial resistance, with differences between the two clades in the rates of resistance evolution and reversion to susceptibility observed for gentamicin, amikacin, colistin, and BL/BLI agents (**Supplemental Table 6**). Moreover, data-driven modeling of rate heterogeneity using *corHMM* also supported the existence of distinct evolutionary rates of resistance across this phylogeny (**Supplemental Table 7**). Lastly, we employed logistic regression to evaluate the association of clade with emergence and spread for each antibiotic. On unadjusted regression, we found that ST258 clade I was positively associated with the following outcomes: amikacin resistance emergence (odds ratio [OR], 2.49; 95% confidence interval [CI], 1.10-5.47), amikacin resistance spread (OR, 9.53; 95% CI, 5.39-17.30), BL/BLI resistance emergence (OR, 2.34; 95 % CI, 1.09-4.99) and BL/BLI resistance spread (OR, 21.19; 95% CI, 5.89-135.76). Conversely, clade II was positively associated with the following outcomes: TMP-SMX resistance spread (OR, 7.50; 95% CI, 4.22-13.91), gentamicin resistance spread (OR, 10.99; 95% CI, 6.01-21.24), and colistin resistance spread (OR, 13.39; 95% CI, 5.72-39.27).

**Figure 3.**
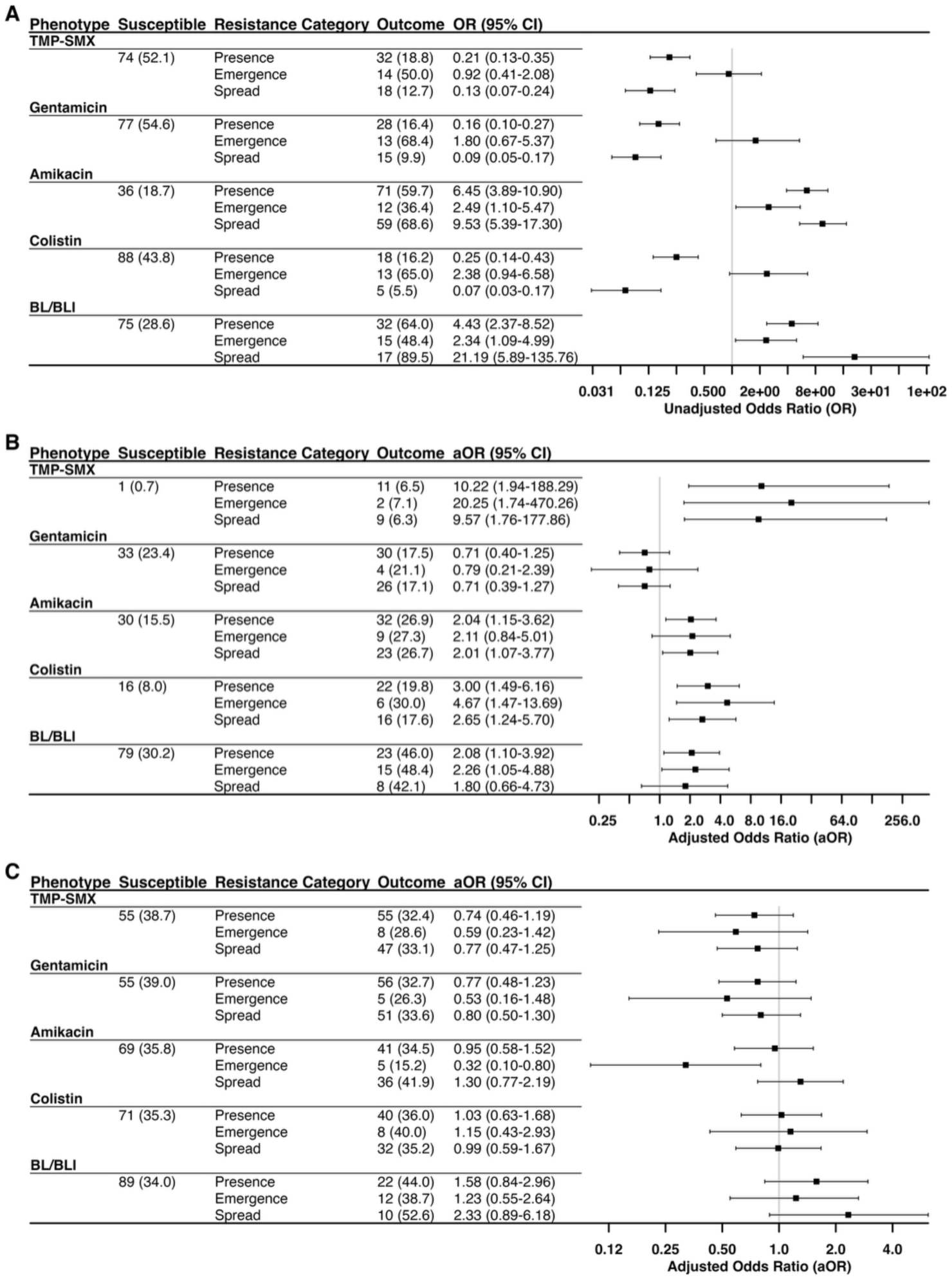
Influence of genetic background and antibiotic exposures on the emergence and spread of resistant lineages **Abbreviations:** BL/BLI, beta-lactam/beta-lactamase inhibitor; CI, confidence interval; OR, odds ratio; TMP-SMX, Trimethoprim-Sulfamethoxazole. Logistic regression was used to analyze the association between our resistance outcomes and (A) having an isolate belonging to clade I of the sequence type 258 phylogeny, (B) exposure to cognate antibiotics, and (C) exposure to dysbiotic antibiotics. For patients with more than one isolate, the first resistant isolate was retained. Each outcome (resistant, emergence, and spread) was compared to susceptible isolates. Exposure to an outcome’s cognate antibiotic was defined as follows: TMP-SMX for TMP-SMX, aminoglycoside exposure for gentamicin and amikacin, polymyxin exposure for colistin, and carbapenem exposure for resistance to BL/BLI agents. Exposure to a dysbiotic agent was defined as a history of exposure to at least one of the following antibiotics: cefepime, ceftaroline, ceftazidime, ceftriaxone, ciprofloxacin, ertapenem, metronidazole, levofloxacin, meropenem, imipenem, and piperacillin/tazobactam [30, 31]. Models for antibiotic exposures were adjusted for patient sex and age at culture collection.

### Phylogenetically informed risk factor analysis revealed shared and unique risk factors associated with the emergence and spread of antibiotic resistance

Having identified cases of resistance emergence and spread, we next explored whether differential risk factors exist across these two groups: phylogenetic singletons (i.e., resistance emergence) and clusters (i.e., spread of circulating resistant lineages). We initially focused on antibiotic use, postulating that exposure to cognate antibiotics would be preferentially associated with selection for resistance emergence. After adjusting for age and sex, cognate antibiotic exposure was positively associated with resistance emergence for TMP-SMX

(adjusted odds ratio [aOR], 20.25; 95% CI, 1.74-470.26), colistin (aOR, 4.67, 95% CI, 1.47-13.69), and BL/BLI agents (aOR, 2.26; 95% CI, 1.05-4.88) (**Figure 3B**). Interestingly, cognate antibiotic exposure was also positively associated with resistance spread for TMP-SMX (aOR, 9.57, 95% CI, 1.76-177.86), colistin (aOR, 2.65, 95% CI, 1.24-5.70), and amikacin (aOR, 2.01, 95% CI, 1.07-3.77). As microbiome-disrupting antibiotics can increase susceptibility to colonization with multidrug-resistant organisms [31], we tested whether these antibiotics were associated with the spread of antibiotic-resistant lineages. No statistically significant, positive association was detected (**Figure 3C**).

To more broadly identify clinical and patient factors that drive the emergence and spread of antibiotic resistance, we performed data-driven multivariable modeling (**Table 1)**. In addition, models were constructed that did not consider phylogenetic context (i.e., phenotypic resistance) to understand how phylogenetic contextualization might provide nuance beyond the standard approach. Inspection of multivariable models revealed risk factors with positive associations for the emergence and spread of antibiotic resistance (**Figure 4**). Cognate antibiotic exposures were detected in both resistance emergence and spread models. TMP-SMX exposure was associated with the emergence and spread of resistance to TMP-SMX. Tobramycin and gentamicin, two aminoglycoside antibiotics, were associated with the emergence and spread of amikacin resistance. Exposure to polymyxin antibiotics were associated with colistin emergence and spread. Increased age was positively associated with the emergence and spread of resistance to BL/BLI agents.

**Figure 4.**
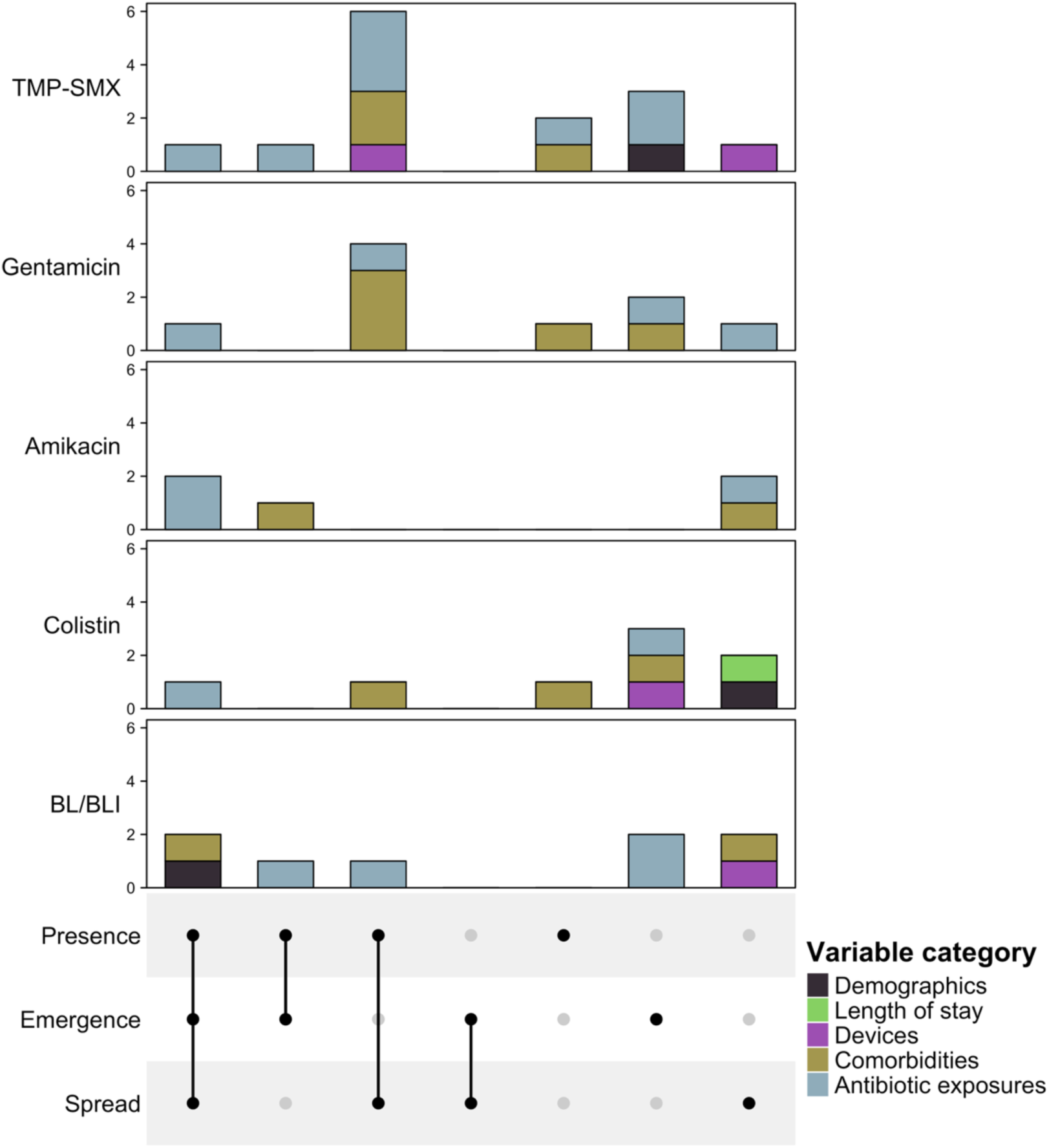
Detection of shared and unique risk factors for the emergence and spread of antibiotic resistance **Abbreviations:** BL/BLI, beta-lactam/beta-lactamase inhibitor; CI, confidence interval; OR, odds ratio; TMP-SMX, Trimethoprim-Sulfamethoxazole. Logistic regression identified risk factors for harboring a resistant strain, harboring a resistant strain inferred to be due to *de novo* evolution, and harboring a resistant strain inferred to be acquired due to cross-transmission. The sharing of model components across these three regression analyses is visualized for resistance to trimethoprim-sulfamethoxazole, gentamicin, amikacin, colistin, and beta-lactam/beta-lactamase inhibitor agents.

**Table 1.** Multivariable logistic regression models reveal differences in risk factors for resistance emergence and spread.

| Phenotype | Multivariable regression models |  |  |  |  |  |  |  |  |
| --- | --- | --- | --- | --- | --- | --- | --- | --- | --- |
| TMP-SMX | Present (n = 170) | OR (95% CI) | p-value | Emergence (n = 28) | OR (95% CI) | p-value | Spread (n = 142) | OR (95% CI) | p-value |
|  | Cephalosporin exposure | 0.40 (0.22-0.72) | 0.0022 | TMP-SMX exposure | 119.13 (5.95-5891.04) | 0.0041 | Acute kidney injury | 0.41 (0.23-0.70) | 0.0014 |
|  | TMP-SMX exposure | 24.97 (4.02-504.50) | 0.0044 | Female sex | 0.35 (0.13-0.86) | 0.0284 | Cephalosporin exposure | 0.40 (0.22-0.73) | 0.0031 |
|  | Acute kidney injury | 0.49 (0.29-0.81) | 0.006 | Tigecycline exposure | 3.66 (1.10-11.75) | 0.0292 | TMP-SMX exposure | 18.93 (3.13-368.12) | 0.0077 |
|  | Gastrostomy tube | 1.86 (1.11-3.18) | 0.0203 | Piperacillin-tazobactam exposure | 0.09 (0.00-0.67) | 0.0709 | Fluoroquinolone exposure | 0.42 (0.20-0.82) | 0.0132 |
|  | Decubitus ulcer stage IV/V | 2.08 (1.12-3.96) | 0.0227 | Polymyxin exposure | 0.10 (0.00-0.80) | 0.0892 | Decubitus ulcer stage IV/V | 2.10 (1.11-4.06) | 0.0247 |
|  | Tigecycline exposure | 2.34 (1.12-5.11) | 0.0278 |  |  |  | Gastrostomy tube | 1.89 (1.09-3.33) | 0.0256 |
|  | Gentamicin exposure | 0.17 (0.02-0.76) | 0.0346 |  |  |  | Gentamicin exposure | 0.09 (0.00-0.54) | 0.0286 |
|  | Amikacin exposure | 0.46 (0.21-0.98) | 0.0465 |  |  |  | Tracheostomy tube | 1.67 (0.94-3.01) | 0.0831 |
|  | Fluoroquinolone exposure | 0.53 (0.28-1.01) | 0.0541 |  |  |  |  |  |  |
|  | Malignancy | 0.52 (0.24-1.08) | 0.0803 |  |  |  |  |  |  |
| Gentamicin | Present (n = 171) | OR (95% CI) | p-value | Emergence (n = 19) | OR (95% CI) | p-value | Spread (n = 152) | OR (95% CI) | p-value |
|  | Decubitus ulcer stage IV/V | 2.23 (1.22-4.20) | 0.0105 | Congestive heart failure | 3.61 (1.21-10.75) | 0.0199 | Decubitus ulcer stage IV/V | 2.57 (1.37-4.96) | 0.0038 |
|  | Cephalosporin exposure | 0.49 (0.28-0.84) | 0.0108 | Cephalosporin exposure | 0.13 (0.01-0.67) | 0.0513 | Chronic kidney disease | 0.47 (0.27-0.82) | 0.0082 |
|  | Chronic kidney disease | 0.52 (0.31-0.88) | 0.0152 | Carbapenem exposure | 0.27 (0.06-0.95) | 0.0626 | TMP-SMX exposure | 10.85 (1.96-203.63) | 0.0259 |
|  | TMP-SMX exposure | 12.95 (2.22-249.77) | 0.0193 |  |  |  | Cephalosporin exposure | 0.56 (0.32-0.98) | 0.0438 |
|  | Malignancy | 0.48 (0.22-1.00) | 0.0525 |  |  |  | Acute kidney injury | 0.60 (0.36-1.00) | 0.0501 |
|  | Acute kidney injury | 0.65 (0.39-1.06) | 0.0858 |  |  |  | Gentamicin exposure | 0.23 (0.03-1.01) | 0.0773 |
| Amikacin | Present (n = 119) | OR (95% CI) | p-value | Emergence (n = 33) | OR (95% CI) | p-value | Spread (n = 86) | OR (95% CI) | p-value |
|  | Tobramycin exposure | 2.82 (1.14-7.40) | 0.0279 | Tobramycin exposure | 3.79 (1.03-12.65) | 0.0334 | Cephalosporin exposure | 1.88 (1.06-3.33) | 0.0297 |
|  | Gentamicin exposure | 4.44 (1.24-20.80) | 0.0318 | Gentamicin exposure | 5.61 (0.94-33.04) | 0.0474 | Gentamicin exposure | 4.61 (1.08-23.30) | 0.0424 |
|  | Malignancy | 1.93 (0.96-3.88) | 0.0647 | Malignancy | 2.55 (0.89-6.69) | 0.0649 | COPD or chronic bronchitis | 1.80 (0.96-3.35) | 0.0618 |
|  |  |  |  |  |  |  | Tobramycin exposure | 2.52 (0.88-7.24) | 0.0806 |
| Colistin | Present (n = 111) | OR (95% CI) | p-value | Emergence (n = 20) | OR (95% CI) | p-value | Spread (n = 91) | OR (95% CI) | p-value |
|  | Polymyxin exposure | 3.85 (1.85-8.28) | 4e-04 | Indwelling urinary catheter | 0.15 (0.04-0.49) | 0.0039 | Polymyxin exposure | 3.19 (1.45-7.11) | 0.0039 |
|  | Ventilator-dependent respiratory failure | 0.54 (0.30-0.95) | 0.0353 | Polymyxin exposure | 7.87 (1.92-34.78) | 0.0042 | Underweight or malnourished | 0.47 (0.24-0.87) | 0.0208 |
|  | Underweight or malnourished | 0.53 (0.29-0.95) | 0.0361 | COPD or chronic bronchitis | 4.60 (1.54-14.26) | 0.0064 | Length of stay, 1 day unit | 0.99 (0.98-1.00) | 0.0667 |
|  |  |  |  | Tigecycline exposure | 3.95 (1.11-13.41) | 0.0277 |  |  |  |

| BL/BLI | Present (n = 50) | OR (95% CI) | p-value | Emergence (n = 31) | OR (95% CI) | p-value | Spread (n = 19) | OR (95% CI) | p-value |
| --- | --- | --- | --- | --- | --- | --- | --- | --- | --- |
|  | Age, 1 year unit | 1.04 (1.01-1.06) | 0.0036 | Age, 1 year unit | 1.03 (1.01-1.06) | 0.025 | Cephalosporin exposure | 5.74 (2.03-17.14) | 0.0011 |
|  | Cephalosporin exposure | 2.21 (1.13-4.27) | 0.0186 | Carbapenem exposure | 2.41 (1.08-5.37) | 0.0299 | COPD or chronic bronchitis | 4.16 (1.38-12.54) | 0.0102 |
|  | Carbapenem exposure | 1.94 (1.02-3.69) | 0.0424 | Tigecycline exposure | 2.32 (0.90-5.60) | 0.0683 | Age, 1 year unit | 1.04 (1.00-1.09) | 0.0383 |
|  | Underweight or malnourished | 0.42 (0.16-0.95) | 0.0498 | Linezolid exposure | 0.17 (0.01-0.87) | 0.0894 | Indwelling urinary catheter | 3.10 (1.07-10.49) | 0.0478 |
|  |  |  |  | Underweight or malnourished | 0.39 (0.11-1.06) | 0.093 | Underweight or malnourished | 0.29 (0.06-1.00) | 0.0744 |

**Abbreviations:** BL/BLI, beta-lactam/beta-lactamase inhibitor; CI, confidence interval; COPD, chronic obstructive pulmonary disease; OR, odds ratio; TMP-SMX, trimethoprim-sulfamethoxazole

Independent risk factors in resistant models tended to segregate into emergence or spread models, supporting the hypothesis that the standard approach imprecisely merges two distinct populations (**Figure 4**; **Table 1**). Several variables were identified as independent risk factors for the emergence of resistance, but not its spread. Antibiotic exposures were often associated with the emergence of resistance. Tigecycline was associated with emergence of resistance to TMP-SMX, colistin, and BL/BLI agents. Exposure to carbapenems was associated with the emergence of resistance to BL/BLI agents but not spread. Additional risk factors that were unique to resistance emergence were detected, notably associations between congestive heart failure and the emergence of gentamicin resistance and malignancy with the emergence of amikacin resistance.

While models for resistance spread included exposure to antibiotics, additional proxies for complexity of care, notably patient comorbidities and indwelling medical devices, were uniquely associated with resistance spread. The presence of stage IV+ decubitus ulcers was associated with the spread of resistance to TMP-SMX and gentamicin. Chronic respiratory comorbidities, specifically the presence of chronic obstructive pulmonary disorder or chronic bronchitis, were associated with the spread of amikacin and BL/BLI resistance. This comorbidity was also associated with the emergence of colistin resistance. Devices were also often associated with the spread of resistant lineages, with presence of a gastrostomy and tracheostomy associated with the spread of TMP-SMX resistance, while presence of an indwelling urinary catheter was associated with the spread of BL/BLI resistance. Highlighting its potentially complex role in resistance dynamics, cephalosporin exposure was positively associated with spread of amikacin and BL/BLI resistance but negatively associated with spread of TMP-SMX resistance, alongside the emergence and spread of resistance to gentamicin.

Collectively, these analyses suggest that the traditional modeling approaches may mask important differences in the patient characteristics and clinical practices that drive the emergence and spread of antibiotic resistance.

## DISCUSSION

To test the hypothesis that more refined insights into the drivers of antibiotic resistance could be attained by considering the distinct pathways to resistance acquisition, we developed a phylogenetically informed approach, compiled as the easy-to-use, open-source R package *phyloAMR*. This tool enables the partitioning of patients from a densely sampled cohort into those putatively acquiring resistant strains via *de novo* evolution or cross-transmission events. Applying *phyloAMR* to a regional collection of CRKP ST258 revealed differences in the relative roles of emergence and spread for five critical antibiotics. We also identified differential impacts of ST258 clade, antibiotic exposures, and clinical characteristics on the emergence and spread of each antibiotic resistance phenotype.

Modeling the acquisition of antibiotic resistance as a binary state of phenotypic resistance (i.e., resistant or susceptible) oversimplifies and masks complexity. Our phylogenetic characterization revealed considerable differences in the contribution of *de novo* evolution and cross-transmission to resistance proliferation in a regionally disseminating bacterial lineage, CRKP ST258, even among antibiotics with comparable frequencies of resistance. Differences in the dynamics of resistance evolution across these antibiotics may reflect differences in magnitudes of selection, the fitness and transmissibility of resistant strains, or the contribution of within-host dynamics [41]. These findings highlight the potential for whole-genome sequencing and phylogenetics to enhance traditional epidemiological investigations into precise microbial and host-associated factors that influence the evolution and spread of antibiotic resistance. Furthermore, phylogenetic contextualization can be leveraged to inform the development of innovative antimicrobial stewardship and infection prevention strategies to account for the dynamics of resistance evolution and spread to prolong the long-term efficacy of an antibiotic.

Our analysis revealed striking differences in the rates of resistance emergence and spread across clades of epidemic lineage CRKP ST258. Prior *in vitro* experimentation and epidemiological studies have observed that resistance-conferring genotypes can impose varying fitness costs across distinct genetic backgrounds and that genetic background influences the propensity for developing mutational resistance [42–46]. The genetic background’s influence on cross-transmission, as observed with colistin resistance, may also influence the dynamics of resistance evolution [47]. Our analysis also identified clade-specific differences in the dynamics of resistance to meropenem-vaborbactam and imipenem-relebactam, two recently approved BL/BLI agents that were not yet clinically available in this population. Investigations of phylogenetic clustering, like those presented in this manuscript, could inform surveillance efforts by identifying genetic backgrounds with a high propensity for resistance development or spread across healthcare systems, as well as distinguishing microbial risk factors for resistance emergence and spread.

Integrating phylogenetic context into risk factor modeling supports the emerging hypothesis that patient comorbidities and medical practices exert distinct selective pressures for the evolution [48] and cross-transmission of antibiotic-resistant lineages [49, 50]. Like prior studies [3–5], we observed that cognate antibiotic exposure was often an independent risk factor for acquisition of antibiotic resistance, potentially through selective pressure. Our modeling additionally suggests that other non-cognate antibiotic exposures can also differentially drive the spread of antibiotic resistance. While we observed that antibiotic exposures were predominantly identified as risk factors when modeling resistance emergence, distinct antibiotic exposures and markers of clinical severity, notably comorbidities and indwelling medical devices, also contribute to the spread of resistant lineages. Therefore, we postulate that antimicrobial stewardship interventions could be tailored to the specific dynamics for each antibiotic-resistant lineage while also considering the underlying patient population. Leveraging the rich phylogenetic contextualization of antibiotic resistance provides novel insights for development of innovative strategies to detect and prevent the emergence and spread of antibiotic-resistant organisms.

Our study has several limitations. First, our collection only included clinical CRKP isolates, but not those from asymptomatic active surveillance which could have facilitated a more comprehensive view of antibiotic resistance dynamics. This limitation is anticipated to be mitigated due to the LTACH population setting, in which dense culture sampling is frequently performed for clinical evaluation [51, 52]. Second, we analyzed only select clinical factors that may select for antibiotic resistance or increase the susceptibility for acquiring a resistant strain. Future investigations should address the evaluation of additional medical comorbidities and exposure to non-antibiotic medications. Third, our analysis was performed on an LTACH population with a high frequency of antibiotic exposures and use of indwelling devices, which may limit the generalizability of our risk factor analyses. Fourth, our ability to detect statistically significant associations between patient characteristics and our phylogenetically-informed resistance outcomes, notably the emergence of antibiotic resistance, was limited by a small sample size.

In conclusion, whole-genome sequencing and phylogenetic characterization of antibiotic resistance in a densely sampled LTACH cohort improved our understanding of antibiotic resistance emergence and spread across a healthcare network. Applying a phylogenetically-informed approach can generate testable hypotheses regarding the pathways to acquisition of antibiotic resistance and inform the development of targeted interventions to reduce the global burden of resistance and extend the long-term efficacy of antibiotics. Future studies, including additional clinical populations (e.g., acute care hospitals, skilled nursing facilities) and microbial species, should investigate whether phylogenetics can consistently reveal trends concerning how bacteria develop resistance and how resistant bacteria spread to new hosts.

## Data Availability

Data produced in the present study are available upon reasonable request to the authors

## Data availability

Code, results, and select de-identified data for the manuscript can be found at https://github.com/kylegontjes/phylogenetic-resistance-ms.

## SUPPLEMENTAL MATERIAL

**Supplementary Table 1.**
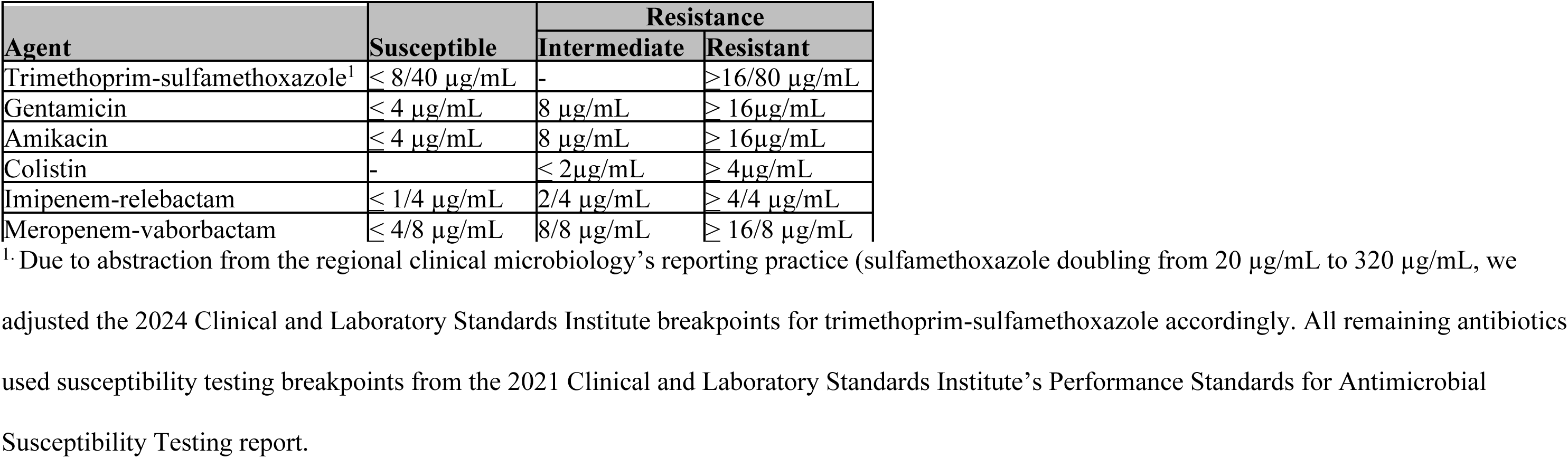
Minimum-inhibitory concentration breakpoints applied in this study.

**Supplemental Table 2.** Variables considered for multivariable logistic regression modeling.

| Variables | Type | Description |
| --- | --- | --- |
| **Demographics** |  |  |
| Age | Continuous | Age in years |
| Sex | Binary | Female or male sex |
| Length of stay | Continuous | Days in the facility preceding isolate collection |
| **Indwelling devices** | Binary | Presence of the device on the day of isolate collection |
| Tracheostomy tube |  |  |
| Central venous catheter |  |  |
| Indwelling urinary catheter |  |  |
| Gastrostomy tube |  |  |
| **Comorbidities** | Binary | Presence of comorbidities on the day of isolate collection |
| Acute kidney injury |  |  |
| Chronic kidney disease |  |  |
| Ventilator-dependent respiratory failure |  |  |
| Underweight or malnourished | | Body mass index $\leq 18.5$ kg/m<sup>2</sup> |
| Congestive heart failure |  |  |
| Decubitus ulcer stage IV/V |  |  |
| COPD or chronic bronchitis |  | Chronic obstructive pulmonary disease or chronic bronchitis |
| Brain injury |  | Anoxic brain injury from a cerebral vascular accident, causing paresis. |
| Malignancy |  |  |
| Obesity | | Body mass index $\geq 30$ kg/m<sup>2</sup> |
| **Antibiotic exposures in the preceding 30 days** | Binary | Exposure of $\geq 1$ day of antibiotics modeled as a binary variable |
| Intravenous vancomycin |  |  |
| Carbapenem |  | Exposure to ertapenem, imipenem, and/or meropenem |
| Cephalosporin |  | Exposures to third-, fourth-, and/or fifth-generation cephalosporins, notably ceftriaxone, ceftazidime, cefepime, or ceftaroline |
| Aminoglycoside |  | Exposures to amikacin, tobramycin, and/or gentamicin. Aminoglycoside exposure was modeled separately, as gentamicin and amikacin resistance had distinct distributions across the phylogenetic tree |
| Metronidazole |  |  |
| Fluoroquinolone |  | Exposure to levofloxacin and/or ciprofloxacin |
| Linezolid |  |  |
| Tigecycline |  |  |
| Piperacillin-tazobactam |  |  |
| Polymyxin |  | Exposure to polymyxin B and/or colistin |
| Daptomycin |  |  |
| Aztreonam |  |  |
| Trimethoprim-sulfamethoxazole |  |  |

**Supplemental Table 3.** Characteristics of the study population at the time of first isolate collection.

| Characteristic | Overall (n=312) |
| --- | --- |
| **General characteristics** |  |
| Age, y, median (IQR) | 72.72 (18.2) |
| Female Sex, no. (%) | 149 (47.8) |
| Length of stay before culture, median (IQR) | 21 (37) |
| **Indwelling device presence** |  |
| Device count, median (IQR) | 2 (2) |
| Any device present | 274 (87.8) |
| Tracheostomy tube | 219 (70.2) |
| Central venous catheter | 180 (57.7) |
| Indwelling urinary catheter | 174 (55.8) |
| Gastronomy Tube | 117 (37.5) |
| **Medical comorbidities** |  |
| Comorbidity count, median (IQR) | 2 (3) |
| Presence of multiple comorbidities | 201 (64.4) |
| Acute kidney injury | 152 (48.7) |
| Chronic kidney disease | 107 (34.3) |
| Ventilator-dependent respiratory failure | 96 (30.8) |
| Underweight or malnourished | 82 (26.3) |
| Congestive heart failure | 65 (20.8) |
| Decubitus ulcer stage IV/V | 65 (20.8) |
| COPD or chronic bronchitis | 61 (19.6) |
| Brain injury | 59 (18.9) |
| Malignancy | 39 (12.5) |
| Obesity | 17 (5.4) |
| **Systemic antibiotic exposure in the preceding 30 days** |  |
| Any antibiotic exposure | 227 (72.8) |
| Intravenous vancomycin | 138 (44.2) |
| Carbapenem | 99 (31.7) |
| Cephalosporin | 79 (25.3) |
| Aminoglycoside | 63 (20.2) |
| Amikacin | 40 (12.8) |
| Tobramycin | 21 (6.7) |
| Gentamicin | 11 (3.5) |
| Metronidazole | 60 (19.2) |
| Fluoroquinolone | 58 (18.6) |
| Linezolid | 46 (14.7) |
| Tigecycline | 42 (13.5) |
| Piperacillin-tazobactam | 42 (13.5) |
| Polymyxin | 37 (11.9) |
| Daptomycin | 12 (3.8) |
| Trimethoprim-sulfamethoxazole | 11 (3.5) |
| Aztreonam | 10 (3.2) |

**Supplemental Table 4.**
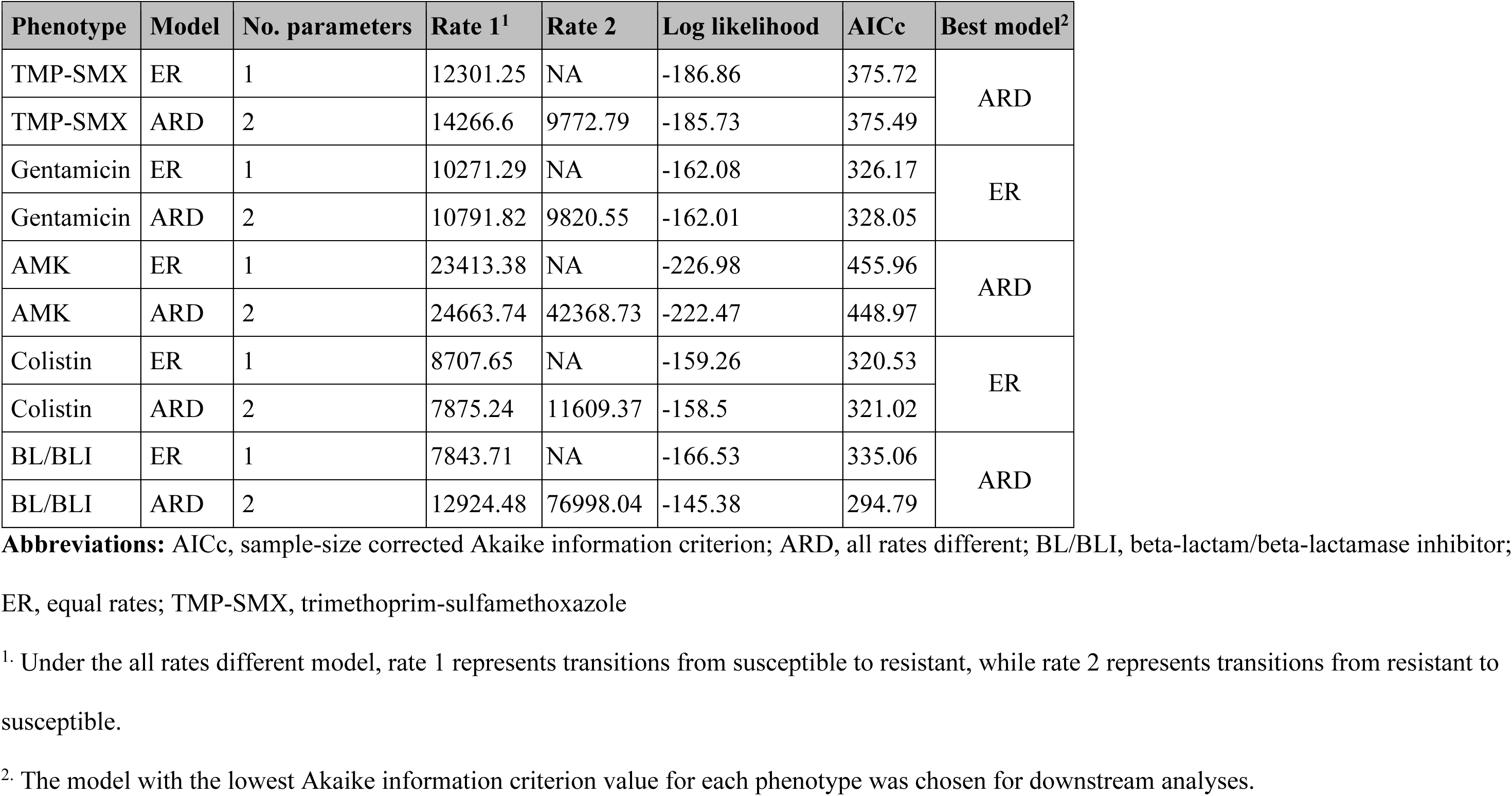
Comparison of ancestral state reconstruction models using Akaike information criteria.

**Supplemental Table 5.** Differences in phylogenetic clustering of antibiotic resistance across clades of *Klebsiella pneumoniae* sequence type 258.

| Phenotype | Clade | Resistant isolates | Resistance frequency | Phylogenetic events | Singletons | Singleton isolates | Clusters | Cluster isolates | Cluster size median (range) | Phylogenetic frequency | Clustering frequency |
| --- | --- | --- | --- | --- | --- | --- | --- | --- | --- | --- | --- |
| TMP-SMX | I | 40 | 29.85% | 19 | 14 | 18 | 5 | 22 | 4 (2-8) | 16.81% | 26.32% |
| TMP-SMX | II | 167 | 66.27% | 18 | 14 | 14 | 4 | 153 | 2.5 (2-146) | 9.33% | 22.22% |
| Gentamicin | I | 36 | 26.87% | 18 | 13 | 17 | 5 | 19 | 3 (2-7) | 11.84% | 27.78% |
| Gentamicin | II | 163 | 64.68% | 9 | 6 | 6 | 3 | 157 | 8 (2-147) | 6.29% | 33.33% |
| Amikacin | I | 79 | 58.96% | 21 | 13 | 14 | 8 | 65 | 7 (2-25) | 7.34% | 38.10% |
| Amikacin | II | 53 | 21.03% | 26 | 21 | 24 | 5 | 29 | 4 (2-15) | 8.58% | 19.23% |
| Colistin | I | 19 | 14.18% | 14 | 13 | 13 | 1 | 6 | 6 (6-6) | 8.92% | 7.14% |
| Colistin | II | 110 | 43.65% | 14 | 7 | 7 | 7 | 103 | 4 (2-71) | 5.83% | 50.00% |
| BL/BLI | I | 37 | 27.61% | 21 | 16 | 17 | 5 | 20 | 2 (2-10) | 15.11% | 23.81% |
| BL/BLI | II | 19 | 7.54% | 17 | 16 | 17 | 1 | 2 | 2 (2-2) | 6.80% | 5.88% |

**Abbreviations:** BL/BLI, beta-lactam/beta-lactamase inhibitor; TMP-SMX, Trimethoprim-Sulfamethoxazole.
<sup>1</sup>. Phylogenetic frequency accounts for the number of episodes a trait occurs across the phylogenetic tree relative to the total number of possible events.
<sup>2</sup>. Clustering frequency characterizes the proportion of phylogenetic events that are phylogenetic clusters.

**Supplemental Table 6.** Evaluation of single-regime and clade-specific evolutionary rate model using *fitmultiMK*.

|  |  | *fitmultiMK* single-regime model |  |  |  | *fitmultiMK* two-regime model |  |  |  |  | Model comparison via the likelihood ratio test |  |  |
| --- | --- | --- | --- | --- | --- | --- | --- | --- | --- | --- | --- | --- | --- |
| Phenotype | Model | Rate 1 | Rate 2 | Log Likelihood | AIC | Clade I Rate 1 | Clade I Rate 2 | Clade II Rate 1 | Clade II Rate 2 | Log Likelihood | AIC | X2 value | p-value |
| TMP-SMX | ER | 12300.44 | NA | -186.86 | 375.71 | 12935.96 | NA | 12016.96 | NA | -186.84 | 377.67 | 0.04 | 0.843 |
| TMP-SMX | ARD | 14277.61 | 9773.99 | -185.75 | 375.49 | 13558.78 | 18657.04 | 18620.48 | 9281 | -184.26 | 376.51 | 2.98 | 0.225 |
| Gentamicin | ER | 10271.16 | NA | -162.08 | 326.16 | 15179.17 | NA | 8494.82 | NA | -160.89 | 325.77 | 2.39 | 0.122 |
| Gentamicin | ARD | 10765.84 | 9852.66 | -162.02 | 328.04 | 18208.26 | 41250.16 | 9512.24 | 8328.1 | -158.65 | 325.29 | 6.75 | 0.034 |
| Amikacin | ER | 23429.23 | NA | -226.98 | 455.95 | 398075.94 | NA | 12357.89 | NA | -212 | 428 | 29.95 | <0.001 |
| Amikacin | ARD | 24665.84 | 42370.76 | -222.47 | 448.94 | 202426.53 | 140691.98 | 16090.46 | 55069.54 | -202.83 | 413.67 | 39.27 | <0.001 |
| Colistin | ER | 8707.39 | NA | -159.26 | 320.52 | 9924.87 | NA | 8150.87 | NA | -159.14 | 322.27 | 0.25 | 0.616 |
| Colistin | ARD | 7889.8 | 11658.15 | -158.52 | 321.04 | 184614.13 | 1097808.65 | 8487.38 | 7764.6 | -146.18 | 300.36 | 24.68 | <0.001 |
| BL/BLI | ER | 7844.06 | NA | -166.53 | 335.05 | 22055.86 | NA | 4480.41 | NA | -158.75 | 321.51 | 15.54 | <0.001 |
| BL/BLI | ARD | 12926.43 | 77010.14 | -145.38 | 294.75 | 45899.85 | 139580.52 | 8497.99 | 82031.48 | -142.39 | 292.77 | 5.98 | 0.05 |

**Abbreviations:** AIC, Akaike information criterion; ARD, all rates different; BL/BLI, beta-lactam/beta-lactamase inhibitor; ER, equal rates; TMP-SMX, Trimethoprim-Sulfamethoxazole.

**Supplemental Table 7.**
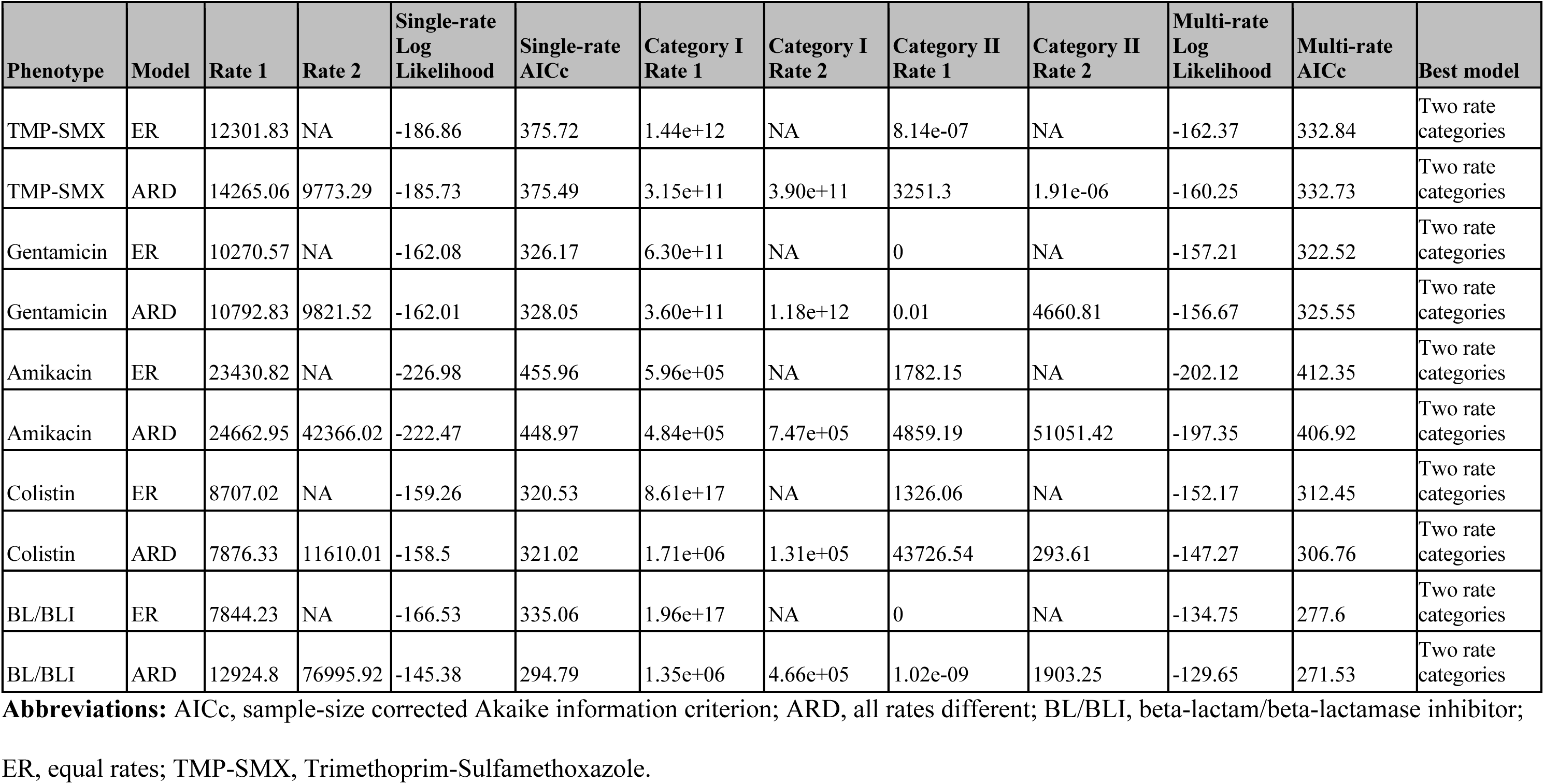
Modeling of single-and multi-rate categories using *corHMM*.

---

**Supplemental Figure 1.**
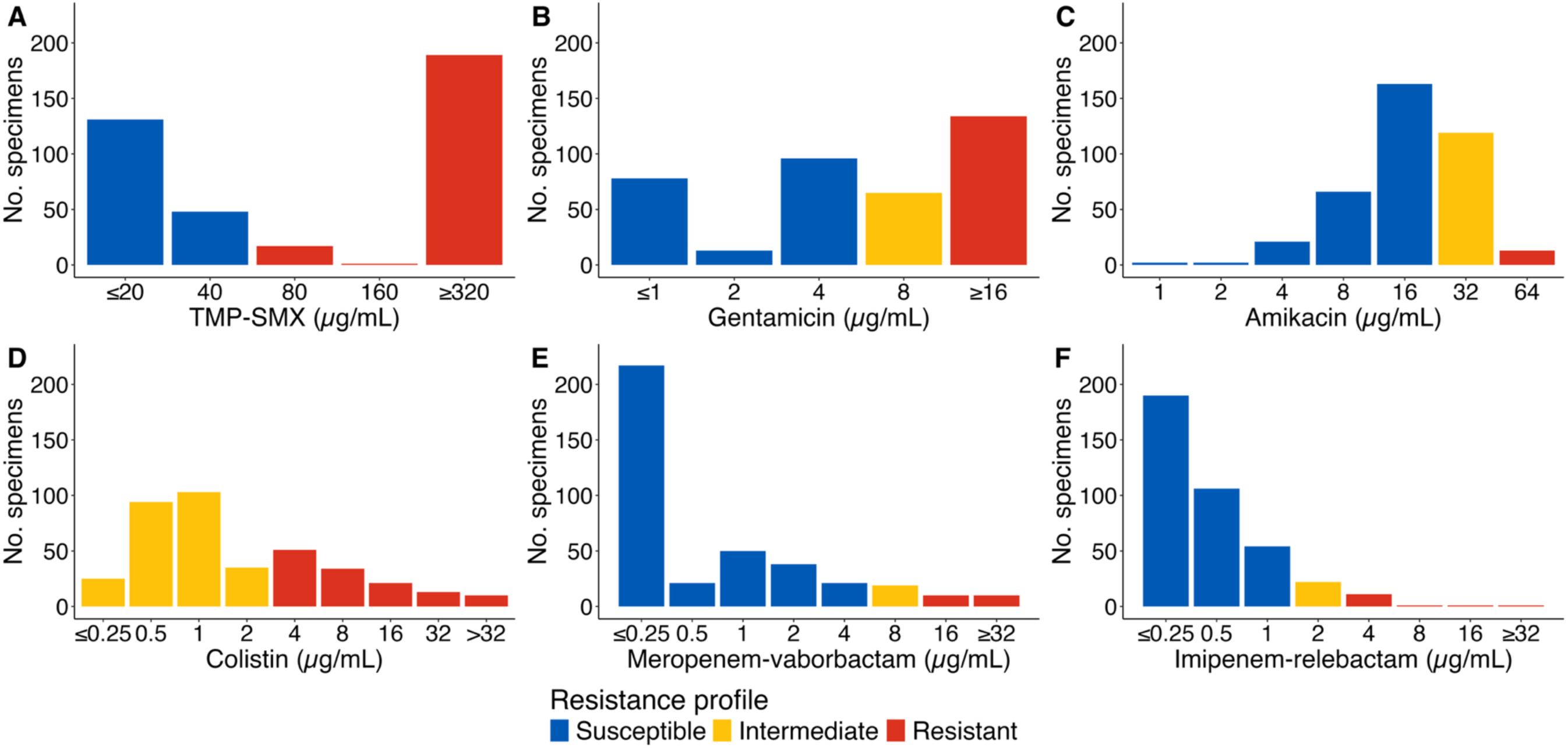
Minimum-inhibitory concentration histogram for each antibiotic of interest **Abbreviations:** TMP-SMX, trimethoprim-sulfamethoxazole

**Supplemental Figure 2.**
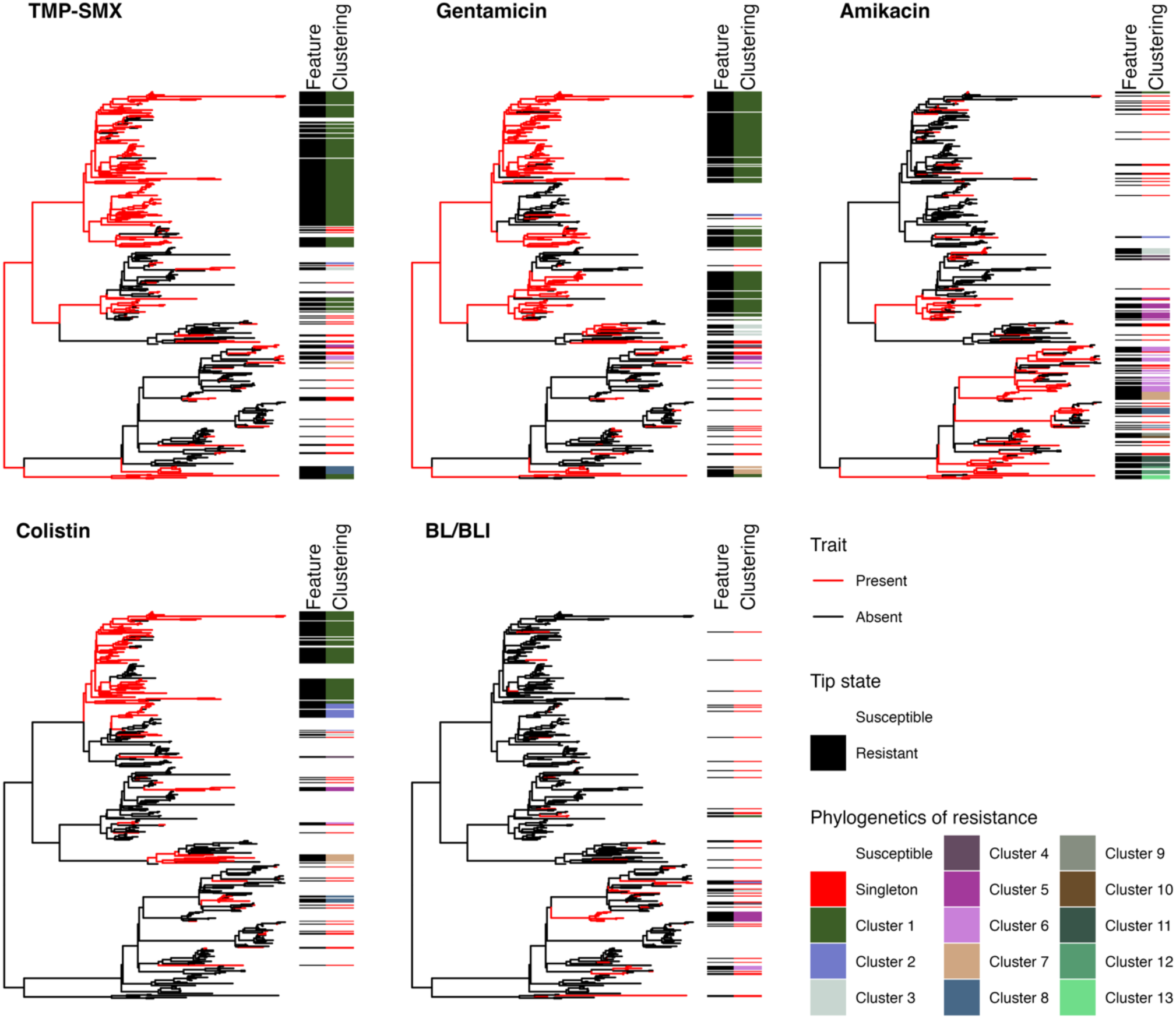
Ancestral state reconstruction and evolutionary history inferences on the phylogenetic tree **Abbreviations:** BL/BLI, beta-lactam/beta-lactamase inhibitors; TMP-SMX, trimethoprim-sulfamethoxazole

